# The Illusion of Understanding: A Randomized Controlled Trial of LLM-Generated Lay Summaries of Brain MRI Reports

**DOI:** 10.64898/2026.08.05.26359773

**Authors:** Bastien Le Guellec, Raphael Bentegeac, Viet-Thi Tran, Marwan El Homsi, Philippe Amouyel, Grégory Kuchcinski, Aghiles Hamroun

## Abstract

**Background:** Large language models have been proposed to improve patient comprehension of radiology reports. However, whether they improve objective understanding remains unproven.

**Purpose:** To evaluate the effect of appending an LLM-generated lay summary to brain MRI reports on objective and subjective patient comprehension in a randomized controlled trial.

**Materials and Methods:** In this randomized controlled trial, 2,727 adult participants from the ComPaRe e-cohort were randomly assigned to interpret six standardized brain MRI reports for headache, presented either in their native format (control; n = 1,401) or appended with a lay summary generated by an open-weights LLM (Mistral Small 3.2) (intervention; n = 1,326). The primary outcome was objective comprehension, defined as the rate of correct classification of whether the report provided a probable explanation for the headache, with ground truth established by four-radiologist consensus. Secondary outcomes included satisfaction, subjective comprehension, anxiety, and willingness to contact a healthcare professional. Generalized estimating equations accounted for repeated within-participant observations.

**Results:** A total of 2,727 participants (mean age, 52 years ± 15; 75.2% women) were evaluated. Objective comprehension did not differ between arms (58.3% vs 59.4%; odds ratio (OR) 0.97; 95% CI: 0.90–1.06; P = .54). The intervention significantly improved overall satisfaction (64.9% vs 36.7%; OR 3.26; 95% CI: 2.93–3.64; P < .001) and subjective comprehension (50.3% vs 24.0%; OR 3.17; 95% CI: 2.82–3.56; P < .001). High anxiety was modestly reduced (25.1% vs 26.6%; OR 0.92; P = .037). The effect on objective comprehension varied by report type (P for interaction < .001): summaries improved comprehension of symptom-explaining reports (42.4% vs 37.4%; P < .001) but reduced it for normal reports (72.5% vs 76.6%; P = .001).

**Conclusion:** LLM-generated lay summaries appended to brain MRI reports improved patient satisfaction and subjective comprehension but did not improve objective comprehension, indicating a gap between perceived and actual understanding that should be addressed before clinical integration.

**Summary Statement:** In this randomized controlled trial of 2,727 participants, LLM-generated lay summaries appended to brain MRI reports improved patient satisfaction and subjective comprehension but did not improve objective comprehension of findings.

**Key Results:**

1. Adding an LLM-generated lay summary to brain MRI reports did not improve the rate of correct identification of symptom-explaining findings (58.3% vs 59.4%; odds ratio, 0.97; 95%CI: 0.90–1.06; P = .54).
2. The intervention markedly improved overall satisfaction (64.9% vs 36.7%; odds ratio, 3.26; 95% CI: 2.93–3.64; P < .001) and subjective comprehension (50.3% vs 24.0%; odds ratio,3.17; 95% CI: 2.82–3.56; P < .001).
3. The effect on objective comprehension varied by report type: summaries improved comprehension of symptom-explaining reports (42.4% vs 37.4%; P < .001) but reduced comprehension of normal reports (72.5% vs 76.6%; P = .001).

## Introduction

The implementation of the 21st Century Cures Act orders the immediate release of clinical data, ensuring that imaging findings are accessible to patients within hours of being signed off(1). Yet radiology reports are characterized by technical jargon, complex sentence structures, and implicit clinical reasoning that presupposes a level of domain knowledge most patients do not possess(2). This mismatch can result in anxiety, inappropriate reassurance, or misguided health decisions.

The inadequate objective comprehension of radiology reports is well documented. Correctly classifying a report as normal or abnormal is achieved at near-chance level when patients read radiology reports unaided(3,4). Beyond the normal/abnormal distinction, correctly identifying the clinical significance of a finding represents an additional layer of complexity, which has received little attention(4).

Brain MRI for headache represents a high-volume clinical scenario in which these communication challenges are particularly acute. Headache accounts for 3.8 million emergency department visits annually in the United States, with neuroimaging utilization reaching 34.8% of cases by 2014, nearly double the rate from 2006(5). Yet the diagnostic yield is low: only 20% of scans reveal findings potentially explaining headache, while incidental findings unrelated to symptoms occur in 22% of examinations(6,7). This environment creates a significant communication gap, as patients may struggle to navigate the subtle semantic space differentiating incidental results from critical findings that truly explain their symptoms(4).

Large language models (LLMs) have emerged as a scalable solution to this problem(8–11). By automatically translating radiology reports in plain-language, they offer the prospect of bridging the gap between clinical language and patient understanding(8). Prior work has demonstrated that reports simplified by LLMs may improve patient satisfaction and perceived comprehension of radiology reports(8,9). However, evidence on whether these subjective gains translate into improved objective comprehension is still lacking (8,12).

The CLEAR-HEAD randomized controlled trial evaluated the effect of adding a lay summary generated by an open-weights LLM on objective and subjective comprehension of brain MRI reports.

## Methods

### Study Design and Registration

This superiority randomized controlled trial was approved by the institutional review board (CSE 2025-3783) and registered at ClinicalTrials.gov (NCT07310394) on November 28th 2025 prior to participant enrollment. It is reported in accordance with the CONSORT 2025 reporting guideline and TRIPOD-LLM(13) checklist.

### Participants

All adult (over 18 years old) participants from the ComPaRe(14) active at the time of invitation (i.e., they connected at least once during the last year) were eligible to participate, with no exclusion criteria applied. The trial was conducted entirely online through the ComPaRe digital platform between April 1st and April 30th 2026.

### Randomization and masking

Eligible participants were randomly assigned by the ComPaRe platform to one of six questionnaire versions using centralized, computer-generated allocation at the point of invitation. The six versions corresponded to three control sub-versions (native reports only) and three intervention sub-versions (reports with LLM-generated lay summary), each presenting the six reports in a different order. Upon their first connection to the questionnaire, participants provided electronic informed consent prior to accessing any study content.

Participants were blinded from the objective of the study. Participants from the intervention arm were not aware that the summaries were generated by a LLM. Outcome assessment was automated and therefore blinded by design. Investigators were blinded to arm assignment during data analysis.

### Intervention

All participants were asked to interpret six anonymized brain MRI reports from patients presenting to the emergency department for headache. The reports were randomly selected from an institutional retrospective cohort(15), purposely sampled to include two normal reports, two reports with symptom-explaining findings, and two reports with incidental findings. Reports were written in French and included the following sections: “Indication”, “Technique”, “Findings” and “Impression”. Ground truth on the normal, incidental or symptom-explaining nature of the findings for each report was established by consensus among four radiologists prior to the study(15). Participants were randomized to read either the native reports alone (control) or the native reports appended with an LLM-generated lay summary (intervention). Summaries were generated prior to the study using Mistral Small 3.2, using the model hyperparameters temperature 0 for a deterministic output, from a zero-shot prompt previously validated by three neuroradiologists (16)(Supplementary Material), and remained identical across all participants exposed to a given report. Because the selected reports were drawn from an internal institutional database, there was no risk of data leakage into the LLM’s pre-training corpus. No concomitant intervention was delivered in either arm. Reports and the lay summaries are available as Supplementary Material.

### Outcomes

The primary endpoint was objective comprehension of the report, defined as the rate of correct answers to the question: “Is a probable explanation for the headache found in this report?” (binary: Yes/No), as pre-registered. Ground truth was obtained from consensus from 4 neuroradiologists, as previously detailed (15). Secondary endpoints were: correct identification of the presence or absence of an anomaly (normal vs. abnormal classification); overall satisfaction; subjective comprehension; willingness to contact a healthcare professional; and self-reported anxiety. Satisfaction, subjective comprehension, and anxiety were assessed on 5-point Likert scales. The remaining secondary endpoints were binary. No harms were anticipated or recorded in this online study involving reading of anonymized reports. The detailed questionnaire is available in Supplementary Material. A first pilot wave involving 50 participants was conducted prior to the main study. Feedback was collected and no significant change to the protocol was made as a result.

### Statistical Analysis

Analysis followed the intention-to-treat principle, including all participants who were randomized and provided electronic informed consent, regardless of the number of reports subsequently completed.

All outcomes were modeled using Generalized Estimating Equations (GEE) with an exchangeable within-participant correlation structure to account for the six repeated observations per participant. Report status (normal/incidental/symptoms-explaining) was included as a fixed effect in all models. The primary outcome was analyzed with a binomial family (results expressed as odds ratios with 95% CI); Likert-scale outcomes with a Gaussian family (results expressed as regression coefficients with 95% CI). Missing responses at the item level were excluded from the relevant model; no imputation was performed, as missingness was minimal (<0.1% of observations). Pre-specified subgroup analyses were performed by sex, age group, educational level, and prior headache history. Effect modification was assessed with Wald chi-squared interaction tests, and these analyses were distinguished from any post-hoc comparisons. Multiple comparisons were corrected using the Benjamini-Hochberg false discovery rate procedure.

No formal sample size calculation was performed. All active members of the ComPaRe e-cohort at the time of invitation were eligible, yielding a convenience sample of 2,727 participants. Post hoc, this sample provided >99% power to detect an absolute difference of 5 percentage points in the primary outcome (two-sided α = 0.05), accounting for the within-participant correlation structure.

All tests were two-sided with a significance threshold of 0.05. Analyses were conducted in Python 3.12 (statsmodels 0.14).

## Results

### Patients characteristics

From April 1st, 2026, to April 30th, 2026, a total of 2,727 patients recruited from the ComPaRe cohort, were randomly assigned to either interpret six anonymized brain MRI reports from patients presenting to the emergency department for headache, with (intervention arm, n = 1,326) or without (control arm, n = 1,401) an LLM-generated lay summary generated using Mistral Small 3.2 (Figure 1).

**Figure 1.**
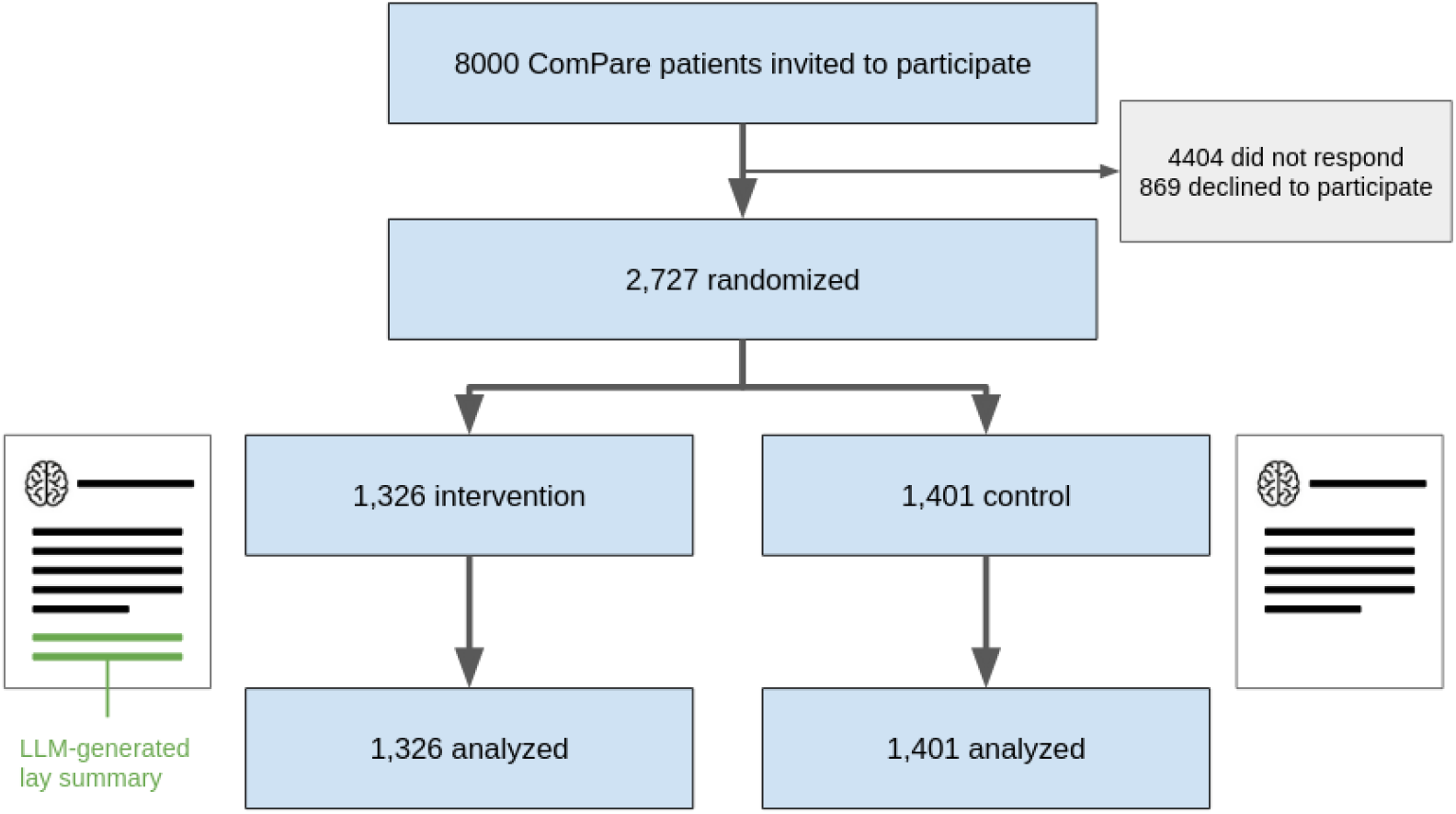
Flow chart.

At baseline, the characteristics of participants were well-balanced between groups (Table 1). The mean age was 51.7 ± 14.8 years in the intervention arm and 51.6 ± 14.8 years in the control arm. Most participants were female (76.0% vs 74.4%) and held a university-level degree (College degree and above: 75.2% vs.73.9%). The proportions with a prior history of headache requiring imaging (32.9% vs. 31.3%) as well as other comorbidities were well balanced.

**Table 1.** Patients characteristics.

| Characteristic | Control<br>(n = 1,401) | Intervention<br>(n = 1,326) |
| --- | --- | --- |
| Age, mean ± SD, y | 51.6 ± 14.8 | 51.7 ± 14.8 |
| **Age group, No. (%)** |  |  |
| 18–40 y | 374 (26.7) | 348 (26.2) |
| 41–55 y | 437 (31.2) | 428 (32.3) |
| 56–70 y | 444 (31.7) | 410 (30.9) |
| >70 y | 146 (10.4) | 140 (10.6) |
| Female sex, No. (%) | 1,042 (74.4) | 1,008 (76.0) |
| **Education, No. (%)** |  |  |
| High school diploma or less | 363 (25.9) | 327 (24.7) |
| College degree or above | 1,036 (73.9) | 997 (75.2) |
| Prior headache requiring neuroimaging, No. (%) | 438 (31.3) | 436 (32.9) |
| **Disease category, No. (%)** |  |  |
| Psychiatry & Mental Health | 467 (33.3) | 421 (31.7) |
| Chronic Pain & Fatigue | 348 (24.8) | 316 (23.8) |
| Neurology | 204 (14.6) | 183 (13.8) |
| Rheumatology & Musculoskeletal | 314 (22.4) | 286 (21.6) |
| Cardiology & Vascular | 258 (18.4) | 262 (19.8) |
| Endocrinology & Metabolism | 302 (21.6) | 307 (23.2) |
| Gastroenterology & Hepatology | 162 (11.6) | 145 (10.9) |
| Pulmonology & Sleep | 213 (15.2) | 207 (15.6) |
| Gynecology & Reproductive Health | 269 (19.2) | 251 (18.9) |
| Urology & Nephrology | 86 (6.1) | 77 (5.8) |
| Oncology | 95 (6.8) | 82 (6.2) |
| Dermatology | 131 (9.4) | 126 (9.5) |
| Ophthalmology | 100 (7.1) | 84 (6.3) |
| ENT & Audiology | 54 (3.9) | 53 (4.0) |
| Infectious Diseases | 159 (11.3) | 145 (10.9) |
| Hematology | 25 (1.8) | 28 (2.1) |
| Immunology & Autoimmune Diseases | 118 (8.4) | 136 (10.3) |
| Rare & Genetic Diseases | 28 (2.0) | 32 (2.4) |

### Primary outcome

The primary outcome was the rate of correct answers to the primary question (“Does this report provide a probable explanation for the headache?”) with ground truth on the normal, incidental or symptom-explaining nature of the findings established by consensus among four radiologists prior to the study(15). This outcome did not differ significantly between groups (58.3% in the intervention arm vs. 59.4% in the control arm; OR = 0.97, 95% CI [0.90–1.06], p = 0.54) (Figure 2).

**Figure 2.**
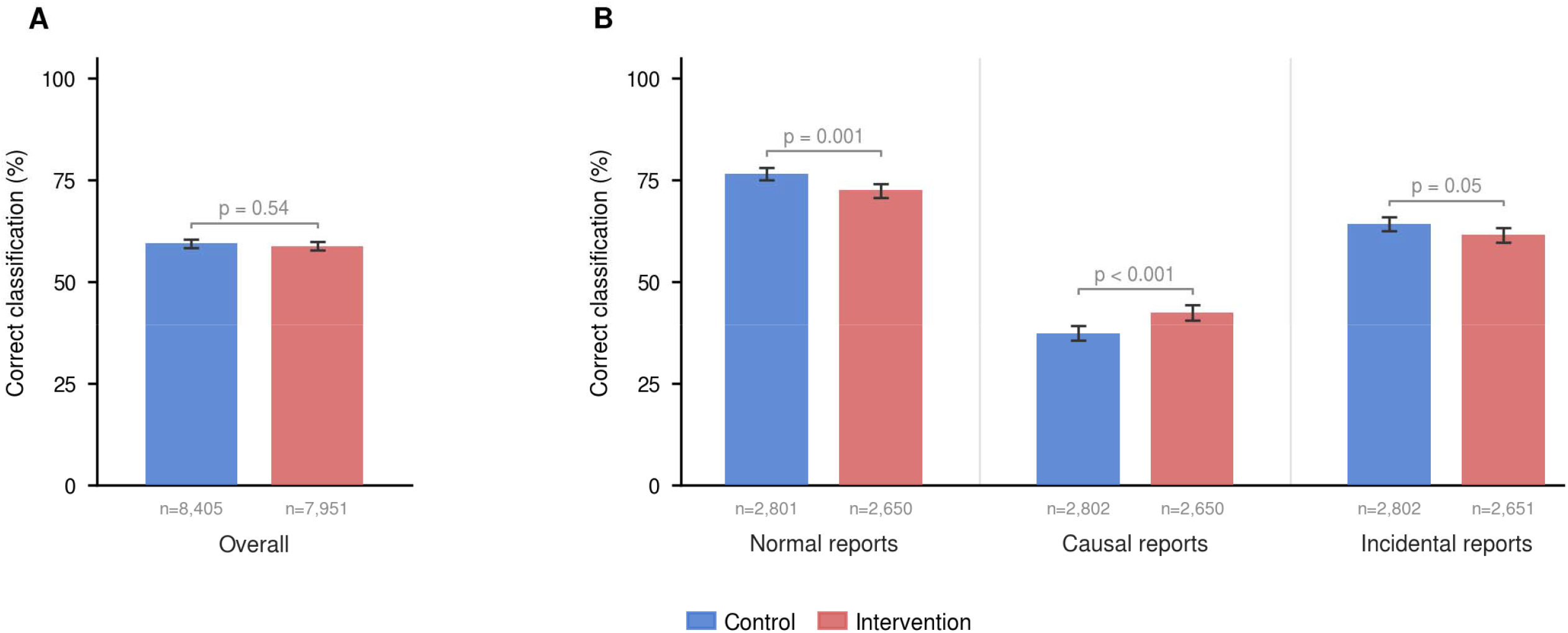
Correct classification of brain MRI reports by study arm, overall and by report type.

The effect of adding an LLM-based lay summary on the primary outcome varied significantly by report type (p for interaction < 0.001). For normal reports, the intervention was associated with a significant reduction in correct classification (72.5% vs. 76.6%; OR = 0.80, 95% CI [0.71–0.92], p = 0.001). In contrast, for reports with symptom-explaining findings, the intervention was associated with a significant improvement in correct classification (42.4% vs. 37.4%; OR = 1.25, 95% CI [1.10– 1.41], p < 0.001). Finally, for incidental-finding reports, the rate of correct answers was marginally lower in the intervention arm (61.5% vs. 64.2%; OR = 0.88, 95% CI [0.78–1.00], p = 0.047). Similarly, the overall rate of correct classification of reports as normal/abnormal did not differ significantly between the intervention and control arms (55.2% vs. 54.1%; OR = 1.02, 95% CI [0.92– 1.14], p = 0.67, Figure 3).

**Figure 3.**
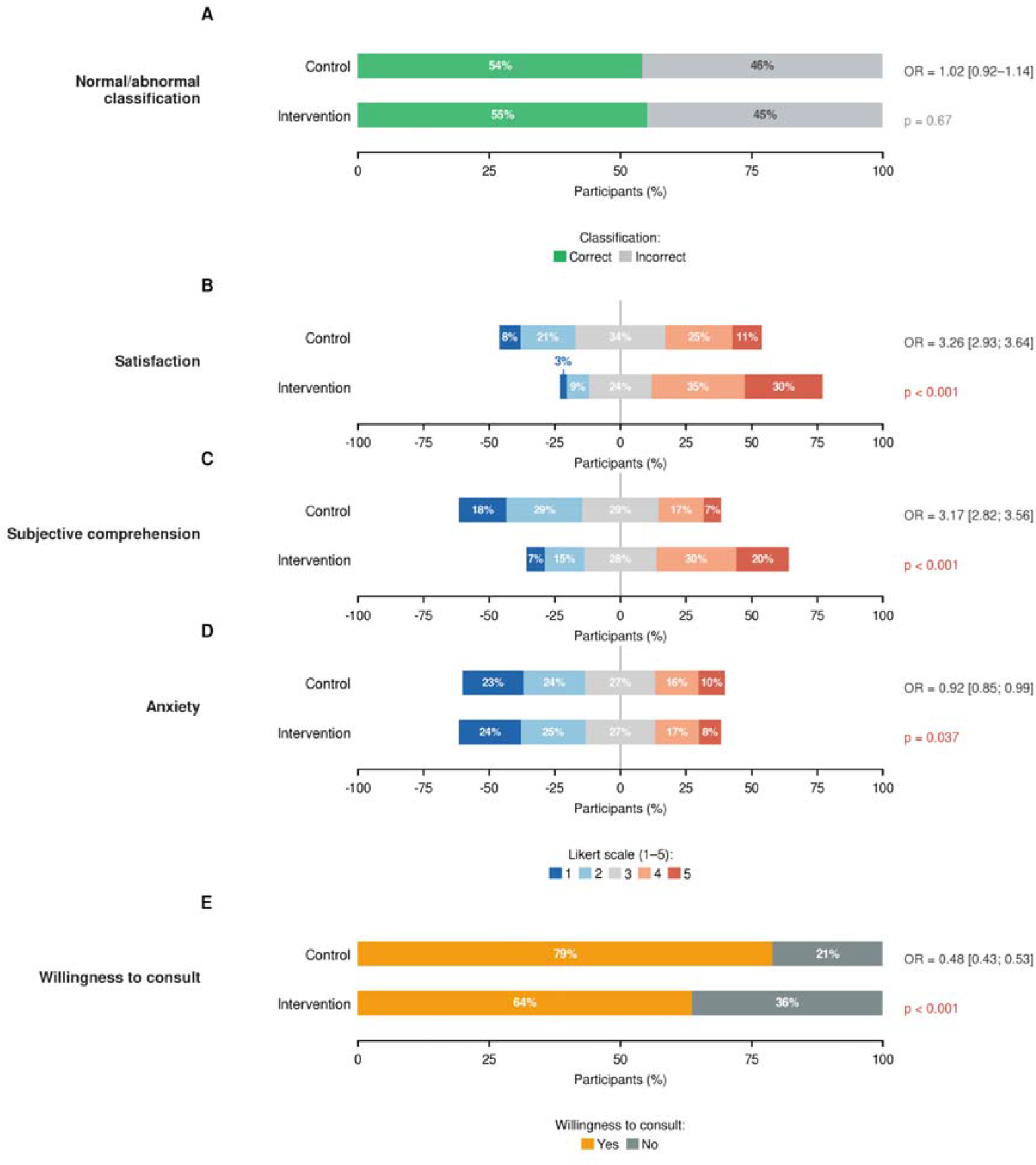
Distribution of secondary patient-reported outcomes by study arm.

### Secondary outcomes

However, we observed significant improvement in all four patient-reported secondary outcomes (Figure 2). Overall satisfaction was markedly higher in the intervention group (64.9% vs. 36.7%; OR

= 3.26, 95% CI [2.93–3.64], p < 0.001, Figure 3). Subjective comprehension improved similarly (50.3% vs. 24.0%; OR = 3.17, 95% CI [2.82–3.56], p < 0.001). The effect on self-reported anxiety was modest but statistically significant, favoring the intervention (25.1% vs. 26.6% reporting high anxiety; OR = 0.92, 95% CI [0.85–0.99], p = 0.037). Participants were less inclined to report a desire to contact a healthcare professional to discuss the results (63.7% vs. 79.0%; OR = 0.48, 95% CI [0.43–0.53], p < 0.001).

The effect of the intervention on the primary outcome was homogeneous across all pre-specified subgroups (Figure 4) with no significant interaction observed for sex, age group, educational level, or prior history of headache requiring neuroimaging. Intervention effects on secondary outcomes were moderated by education level (all pinteraction < 0.05), with the largest gains in satisfaction and comprehension reported by participants with university-level education.

**Figure 4.**
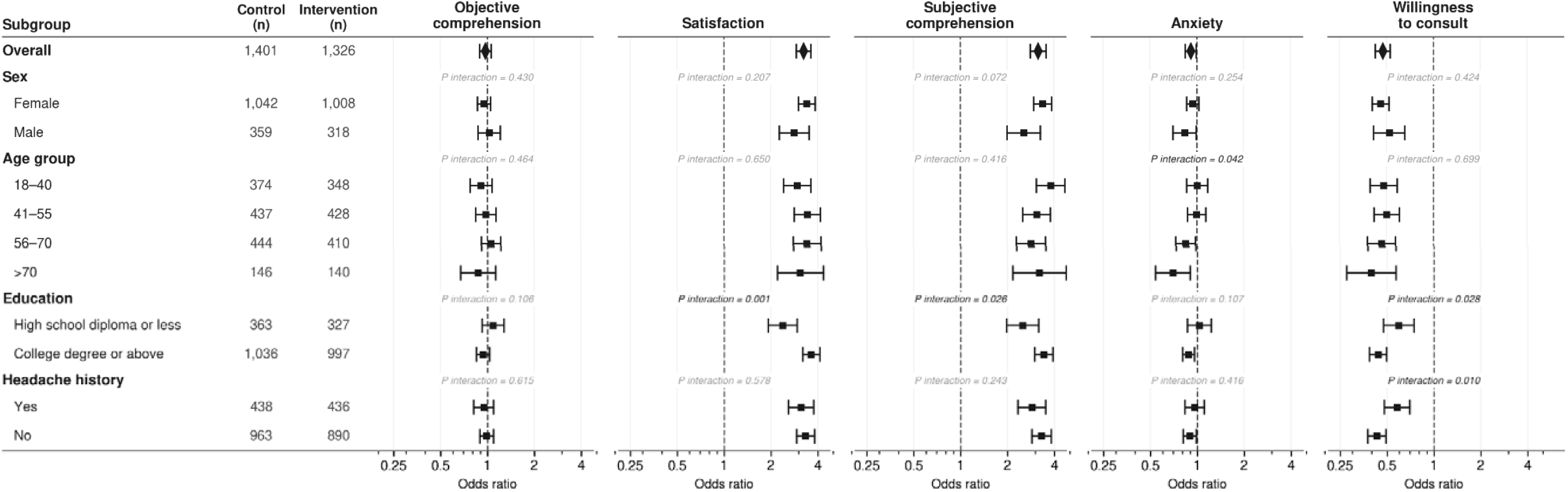
Subgroup analyses for all outcomes.

## Discussion

In this randomized controlled trial, we found that providing LLM-generated patient summaries did not improve objective comprehension of brain MRI reports for headache. However, the intervention yielded significant benefits regarding subjective comprehension, patient satisfaction, and modest anxiety reduction.

The disconnect between the complex vocabulary utilized in radiology reports and the health literacy of the patients reading them is well documented. Cho et al. demonstrated that only one in two patients could successfully comprehend the key message of oncology imaging reports(4). Similarly, Karliner et al. reported that only 51% of patients with an abnormal mammography result understood that their report was actually abnormal(3). Our baseline findings align with this literature, showing an approximate 54% baseline accuracy in correctly classifying MRIs as normal or abnormal, and identifying whether findings were responsible for symptoms.

LLMs hold great promise in deciphering these complex reports. Prucker et al. recently demonstrated a clear improvement in the comprehension of oncology reports translated by LLaMA, an open-source LLM. However, as highlighted in recent reviews(8,17), the true effect of these models on objective comprehension has remained elusive and may not naturally align with perceived comprehension(12,18).

A recent study by Chen et al. on 2,000 readers suggests benefits of translating X-ray reports in plain language on objective comprehension, with a marginal gain of +10% in classification tasks (19).However, their within-subject design comparing original and fully-simplified versions of different reports at the same time and their use of a proprietary model with per-report customized prompts limits their finding. Here, using a strict parallel-arm randomized controlled trial of more complex volumetric studies, we showed no benefit of such an intervention. This discrepancy may reflect a fundamental difference in task complexity. Cognitive load theory distinguishes extraneous load, the burden imposed by jargon and complex syntax, from intrinsic load, which is inherent to the complexity of the content itself(20). Plain-language translation primarily reduces extraneous load, which may suffice for binary normal/abnormal classification of chest X-rays. However, distinguishing incidental from symptom-explaining findings on brain MRI requires integrating clinical context with imaging content, a higher-order reasoning task that carries irreducible intrinsic load. Consistent with this framework, Holderried et al. recently showed that LLM-generated patient letters improved retention of factual, low-complexity information but were far less effective for higher-order comprehension objectives(21).

The paradox of illusory understanding (22) — where explanations are perceived as sufficient by participants but yield variable effects on true comprehension — has been previously shown outside of radiology. Horwitz et al. (23) showed that following discharge instructions, while 98.1% of patients reported understanding the reason for hospitalization, 40.4% failed on objective testing. Two complementary cognitive mechanisms may underlie this dissociation. On the sender side, the “curse of expertise”(24), a bias wherein experts inadvertently overestimate the intelligibility of their explanations, suggests that texts designed by experts may presuppose more background knowledge than patients possess. On the reader’s side, processing fluency(25), the subjective ease with which information is processed, leads readers to equate readability with comprehension. LLM-generated texts, optimized by design for linguistic simplicity(10), may thus inflate perceived understanding without improving actual encoding of clinical content.

This finding carries a dual implication. On one hand, it demonstrates that LLM-generated summaries fulfill a genuine patient need for accessible communication, aligning with the core mandates of the 21st Century Cures Act(1,26). On the other hand, participants in the intervention arm were significantly less inclined to report a desire to contact a healthcare professional (63.7% vs. 79.0%). In the context of unchanged objective comprehension, this reduced drive for clarification may reflect misplaced confidence rather than appropriate reassurance.

The results of our study should not discourage the use of LLMs for radiology report simplification. The positive effect observed for reports containing symptom-explaining findings suggests that LLMs may be effective for specific report types rather than as a universal solution. Future work should explore whether tailoring summaries to report complexity or clinical context can close the gap between perceived and actual comprehension. Promising directions include embedding self-assessment prompts analogous to the teach-back method, which consistently improves objective understanding across clinical settings and enriching summaries with annotated anatomical illustrations, which have been shown to enhance patient information processing beyond text-only modifications. More broadly, recent studies have suggested potential values for other types of reports, including breast and chest imaging(19,27), though those results will need validation with controlled trials.

Our study has several limitations. First, participants read standardized reports rather than their own personal medical records, which may have limited their emotional investment and context(8). This trial design was necessary to isolate the effect of the summary and remove confounding factors such as varying baseline health literacy applied to varying report complexities. Additionally, validation studies have shown that responses to standardized clinical scenarios closely approximate behaviors observed in real-world(28). Second, only a small, curated set of six reports was utilized. However, these were drawn from actual clinical practice, and stratifying them by the nature of their content captures the inherent variability of real-world radiology reports. While this study was not designed to capture all the variance of clinical practice, its controlled conditions establish a baseline for testing the strict effect of the addition of a lay summary. Finally, to maintain homogeneity of the intervention, only a single LLM (Mistral Small 3.2) was tested, limiting the generalizability of the findings to other models or prompting strategies. Nonetheless, utilizing a lightweight, open-weights model ensures a version-controlled solution that is realistically deployable in clinical reading rooms, thereby enhancing the replicability of our results. Furthermore, recent studies indicate that models of similar scale exhibit comparable overall performance in generating patient summaries(16).

In conclusion, we demonstrate that the benefits of LLMs in enhancing patients’ perceived understanding of their medical imaging do not necessarily translate into improved objective comprehension. These findings highlight a critical gap between perceived and actual understanding, driven in part by the very linguistic fluency that makes these tools appealing, and suggest that future implementations should incorporate strategies to calibrate patient confidence against true comprehension.

## Data Availability

Upon request to the corresponding author

